# Task-induced changes in brain entropy

**DOI:** 10.1101/2023.04.28.23289255

**Authors:** Aldo Camargo, Gianpaolo Del Mauro, Ze Wang

## Abstract

Entropy indicates irregularity of a dynamic system with higher entropy indicating higher irregularity and more transit states. In the human brain, regional entropy has been increasingly assessed using resting state fMRI. Response of regional entropy to task has been scarcely studied. The purpose of this study is to characterize task-induced regional brain entropy (BEN) alterations using the large Human Connectome Project (HCP) data. To control the potential modulation by the block-design, BEN of task-fMRI was calculated from the fMRI images acquired during the task conditions only and then compared to BEN of rsfMRI. Compared to resting state, task-performance unanimously induced BEN reduction in the peripheral cortical area including both the task activated regions and task non-specific regions such as the task negative area and BEN increase in the centric part of the sensorimotor and perception networks. Task control condition showed large residual task effects. After controlling the task non-specific effects using the control BEN vs task BEN comparison, regional BEN showed task specific effects in target regions.

## Introduction

Entropy indicates the irregularity or uncertainty of a dynamic system (Clausius, 1865). Statistically, entropy can be defined by the number of states that a system can be configured (Bolzmann, L, 1964) or the information capacity of the dynamic system (Shannon, C.E., 1948). In theory, entropy tends to increase with time and extremely high entropy will destroy the organization of a system. Living organisms are characterized by being highly self-organized (Bak et al., 1989; Bak, P, 1996; Camazine, S, et al., 2020). Counteracting entropy is then crucial to keeping their functional order and the corresponding functions (Bergström, 1969; Pinker, S, 1997; Singer, 2009). The human brain is one of the most complex and self-organized systems known to our human beings and has a prominent need to maintain entropy. While it remains elusive for how this entropy maintaining operates, monitoring brain entropy (BEN) may open a unique window to observe and evaluate brain function and brain health. Recent years, regional BEN has been increasingly characterized using resting-state fMRI (rsfMRI) (Smith et al., 2014, 2015; Sokunbi et al., 2011, 2013; B. Wang et al., 2017; Wang et al., 2014; Wang, Ze, 2012, 2013). Using rsfMRI from a large cohort of healthy subjects, we have identified normal resting BEN distributions (Wang et al., 2014), and its associations with age, sex, educations, and neurocognitions(Wang, 2021). The neurocognitive correlates of resting BEN was further supported by the correlations between regional BEN and the magnitude of task activations and task de-activations in the corresponding activated and de-activated brain regions(Liandong Lin et al., 2022). The translational value of rest BEN has been implicated by its alterations found in various brain diseases (Li et al., 2016; Lin et al., 2019; Sokunbi et al., 2013; Z. Wang et al., 2017; Xue et al., 2019; Zhou et al., 2016). Importantly, we have demonstrated that regional BEN is modifiable through repetitive transcranial magnetic stimulations (Chang et al., 2018; Song et al., 2019), and medication (Liu et al., 2020).

Thus far, the vast majority of BEN mapping research is based on rsfMRI. Few studies have examined regional BEN changes in response to task activation or deactivations. Based on sensorimotor task fMRI from a small cohort of healthy subjects, we have showed task-induced regional BEN reductions in motor and visual cortex(Wang et al., 2014). Using a subset (n=100) of the Human Connectome Project (HCP) cohort (Van Essen et al., 2013), Nezafati et al. (Nezafati et al., 2020) demonstrated reduced BEN in both cortical and subcortical regions during working memory task. In this study, we aimed to examine task performance-induced voxelwise BEN alterations as compared to resting BEN. The major differences from previous work (Nezafati et al., 2020; Wang et al., 2014) were: First, we used the entire large cohort from the HCP to gain more statistic power; Second, we examined seven different tasks to check the generalizability and specificity of the task-induced BEN changes; Third, previous work calculated BEN using the entire task-fMRI time series. In this paper, we did additional work to calculate task fMRI BEN using the fMRI images extracted at all task conditions. We also compared BEN of resting state fMRI to BEN of task fMRI during the non-task blocks to explicitly see the task performance-induced residual effects on the non-task performing brain state.

## Materials and Methods

### Ethics statement

Data acquisition and sharing have been approved by the HCP parent IRB. Written informed consent forms have been obtained from all subjects before any experiments. This study re-analyzed the HCP data and data Use Terms have been signed and approved by the WU-Minn HCP Consortium. Code is available from https://github.com/zewangnew/BENtbx.

### Data descriptions

1096 healthy young adults (age range: 22 – 37 years old, male/female: 550/656) were identified from the HCP (Van Essen et al. 2013). The mean and the standard deviation of years of education were 14.86±1.82 years. The range of years of education was 11-17 years. Participants with missing data were excluded. Structural MRI, rsfmri, and task-fMRI were acquired with a connectome dedicated 3 Tesla (3T) Siemens MR scanner. Detailed protocols used to acquire MRI data for the HCP project are described at https://www.humanconnectome.org and in the literature (Van Essen et al., 2013).

### Task-fMRI

Seven task-fMRI experiments were considered in this study, including motor, social, relational, working memory (WM), gambling, language, and emotion. Detailed task design and imaging parameters can be found in (Barch et al., 2013). To avoid an exhaustive analysis and presentation for the large number of task conditions (each condition consists of a few blocks for a specific task) included in the seven tasks: 1) motor task fMRI: right-hand (RH), left-hand (LH), right-foot (RF), left-foot (LF), and tongue; 2) social task-fMRI: social interaction and random motion; 3) relational task-fMRI: match and relation; 4) WM-fMRI: 0-bk and 2-bk; 5) gambling task fMRI: loss and win condition; 6) language task-fMRI: math and story; 7) emotion task-fMRI: the face condition and shape condition. All tasks were presented to the participants in text or video projected to the screen in the back of the MR scanner. Participants saw the screen through the mirror hooked on the head coil. During the motor task-fMRI scan, participants were instructed through visual cues to either squeeze their left or right toes, or tap left or right fingers, or move their tongue. In the social cognition task, subjects were shown short video clips of objects (squares, circles, triangles) that either interact in some way, or moved randomly on the screen. After the video clip the participants were asked to judge whether the objects had a mental interaction, not sure, or no interaction. During the relational task, participants watched 6 different shapes filled with 1 or 6 different textures. For the relational task condition, the participants had to check whether the shape of texture differs between the objects shown on the screen. For the match task condition, two objects were shown in the top of the screen and one in the bottom. The participants were asked to match the bottom object with the top ones in “shape” or “texture”. WM task consists of several blocks of trials showing pictures of places, tools, faces, and body parts (non-mutilated parts of bodies with no “nudity”). Within each run, the 4 different stimuli were presented in separate blocks. ½ of the blocks used a 2-back working memory task and ½ used a 0-back working memory task as the control condition. The gambling task was a card game, where subjects had to guess the number of the mystery card to win or lose money. The language tasks (Binder et al., 2011), consist of two runs of 4 blocks of math and 4 blocks of story. For the math blocks, participants were asked to answer simple arithmetic questions like five + twelve. The story blocks consist of brief auditory stories, followed by a 2-alternative choice about the topic of the story. The emotional tasks contained blocks of trials asking participants to match the face, or one of the two shapes presented on the screen.

### BEN mapping

BEN maps were calculated for both rsfMRI and task-fMRI using BENtbx (Wang et al., 2014). The preprocessed rsfMRI and task-fMRI provided by HCP were used. BENtbx was updated to be compatible with the CIFTI image format used by HCP and be able to automatically locate the specific task-block associated images based on the event timing file (the EV file). The updated version is available in https://github.com/zewangnew/BENtbx. For each task-fMRI, images from all task blocks were extracted and time concatenated to form a new time series for calculating BEN. For the simplicity of descriptions, the corresponding BEN was named as task BEN; BEN of all non-task performing blocks (the control blocks) was called control BEN; BEN of rsfMRI was called resting BEN or rest BEN. Whenever a task BEN is compared to resting BEN, the rsfMRI time series was truncated to have the same time points as that of the task fMRI for calculating a new resting BEN map. This step was to prevent any potential systematic entropy value difference caused by the difference of time series length and was performed for each task fMRI condition separately.

### Statistical Analysis

A series of group level BEN comparisons were made between the entire task performing conditions and the control condition (non-task blocks), and between the entire task conditions and the resting state, and between the control condition and resting state. The comparison was made using paired t-test as implemented in SPM12. As we described above, for each paired comparison, the two fMRI time series used to calculate BEN were length-matched by truncating the longer time series to have the same length as the shorter one.

Three types of comparisons were made: 1) Rest BEN vs all-task BEN where rest BEN means BEN calculated from the rsfMRI and all-task means fMRI from all task blocks of a task fMRI experiment. Calculating BEN from the task blocks only was to avoid the block-design induced systematic entropy reduction. The rest BEN – all-task BEN comparison was to assess the task-induced BEN changes compared to the resting state; 2) control BEN vs all-task BEN where control BEN was calculated from the non-task performing blocks (the task control blocks). Similar to the standard task activation detection in task fMRI analysis, this comparison was to examine the within-session task-induced BEN changes; 3) resting BEN vs control BEN comparison for examining the residual task effects in the control condition of the task fMRI scan. All comparisons were repeated for every task fMRI experiment. BEN changes were defined by a statistic threshold of t>5.7 (corresponding to p<0.001 with multiple comparison correction through the family-wise error (FWE) method (Nichols & Hayasaka, 2003)).

## Results

Fig 1 shows the statistical analysis results of BEN comparisons between resting state, the task fMRI control state, and the motor task performing state. As compared to both the resting state (Fig. 1A) and the within-session control state (the non-task performing state, Fig. 1B), the visual instruction guided motor task performance reduced entropy in the peripheral brain regions in visual cortex, the superior section of the motor cortex (face, nose, mouth, and pharynx etc), temporal cortex, and prefrontal cortex (lateral and medial orbito-frontal cortex (OFC), and the dorso-lateral prefrontal cortex (DLPFC)) but increased entropy in the more interior part of brain cortex and subcortical areas such as the superior part of motor network (including the foot and hand area, the supplement motor area (SMA), and precentral cortex), cerebellum, fusiform, the limbic area (including hippocampus, amygdala, ventral striatum, basal ganglia, insula, thalamus, and cingulate), and posterior cingulate cortex (PCC) and precuneus. Compared to the resting state, the task control state (Fig. 1C) showed similar BEN reduction and increase patterns as in Fig. 1A.

**Figure 1.**
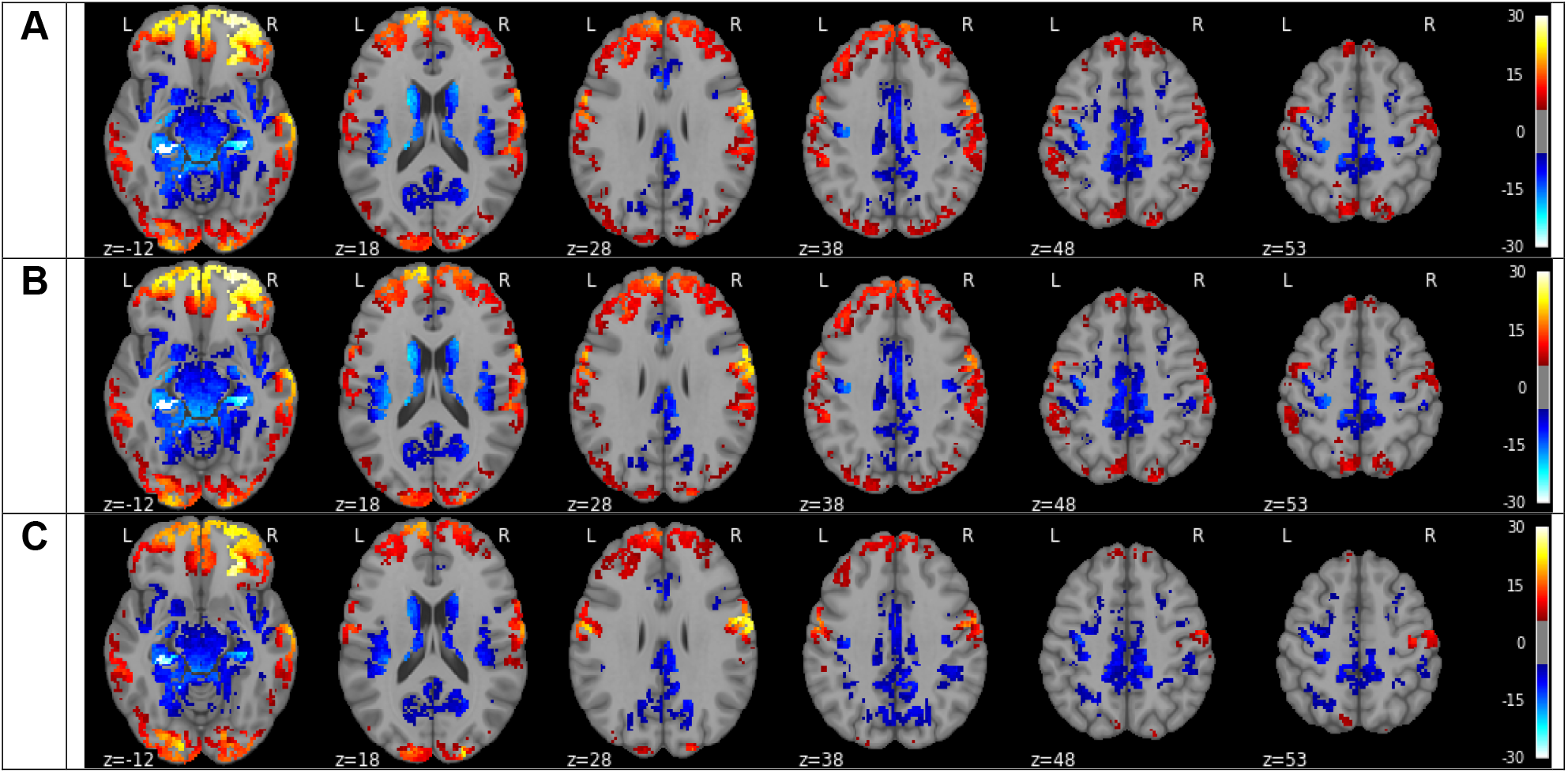
Statistical difference between resting BEN (BEN of rsfMRI), control BEN (BEN of the control condition of the fMRI scan), and all-task BEN (BEN of fMRI images from all task conditions) of the motor task fMRI. A) Resting BEN minus all-task BEN, B) control BEN vs all-task BEN; C) resting BEN minus control BEN. Red/blue means higher/lower entropy in the first state of the comparison (such as the resting state in the resting BEN minus all-task BEN comparison), respectively.

Fig 2 is the statistical BEN comparison results for the social task fMRI. The resting vs social task BEN comparison results (Fig. 2A) showed similar peripheral-interior decrease and increase bifurcation patterns to those shown in the resting BEN vs motor task BEN comparison results (Fig. 1A). The major difference was that social task induced spatially more spread BEN reduction in dorso-lateral prefrontal cortex (DLPFC) and the interior part of angular gyrus, which both are part of the dorsal attention network (DAN). Meanwhile, social task increased BEN in anterior visual cortex and the central part of the sensorimotor system (cerebellum, striatum, hippocampus, basal ganglia, thalamus, superior motor cortex, and SMA). Compared to the task control condition (Fig. 2B), social task only showed reduced BEN in visual cortex, fronto-parietal network and motor cortex. Compared to the resting state, the task control condition (Fig. 2C) showed similar entropy reduction and increase as Fig 2A, with some differences in the visual cortex and temporal lobe.

**Figure 2:**
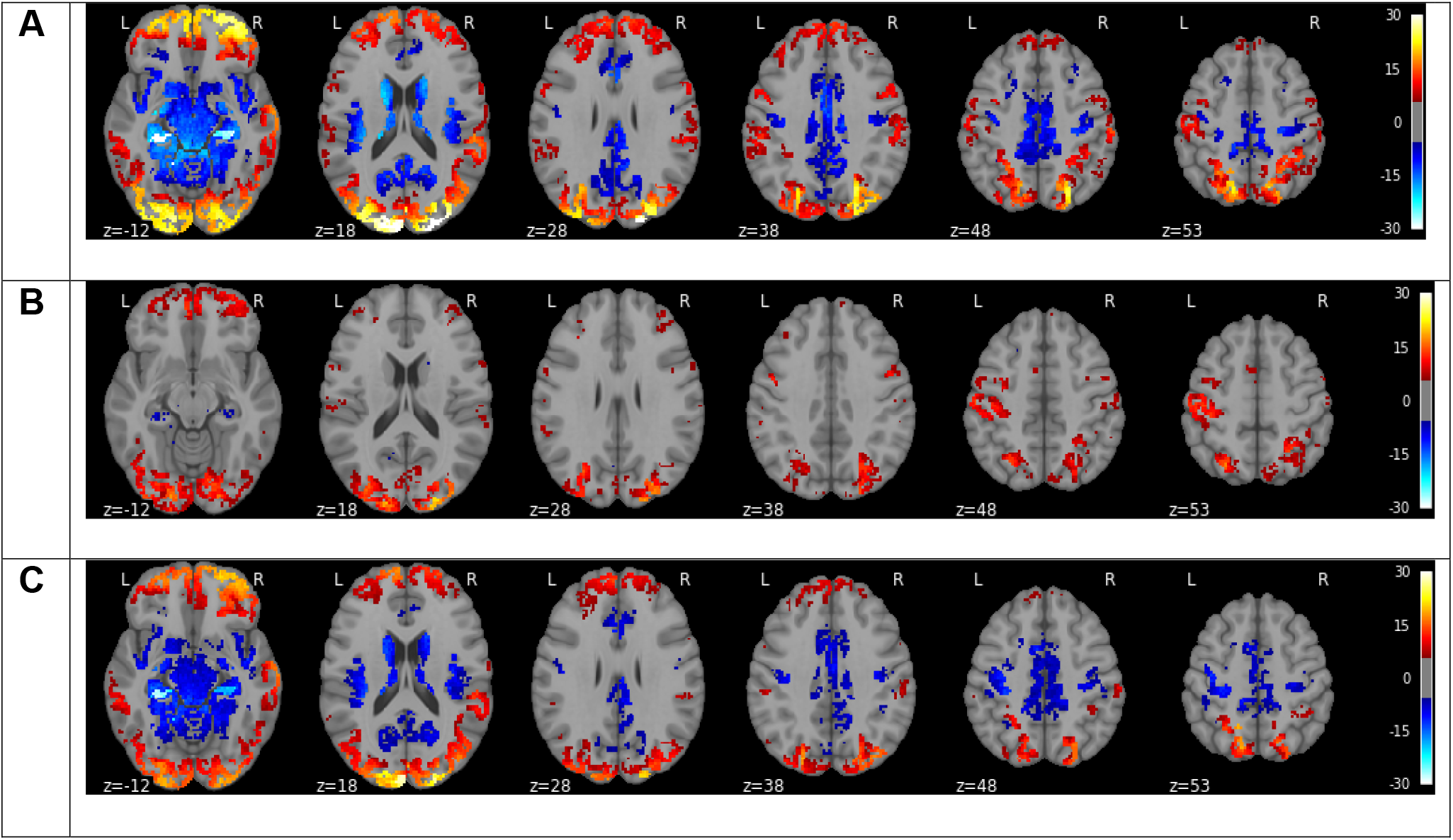
Regional BEN changes invoked by social task performance. A) resting BEN vs BEN of the task social; B) control BEN vs BEN of the task social condition; C) resting BEN minus control BEN (BEN of the non-task condition). Hot and blue color mean higher and lower BEN at the resting state, respectively. Color bars indicate the display window for the t-value of the paired-t test.

Figure 3 shows the statistical comparison results of the relational task induced BEN changes. The three comparison results (Figs 3A, 3B, 3C) are quite similar to those shown in Fig. 2 (the social task). The biggest BEN change patterns were in the resting vs control comparison (Fig. 3C). Compared to resting state, the control condition for the relational task showed (Fig. 3C) less BEN reduction in the parietal cortex.

**Figure 3:**
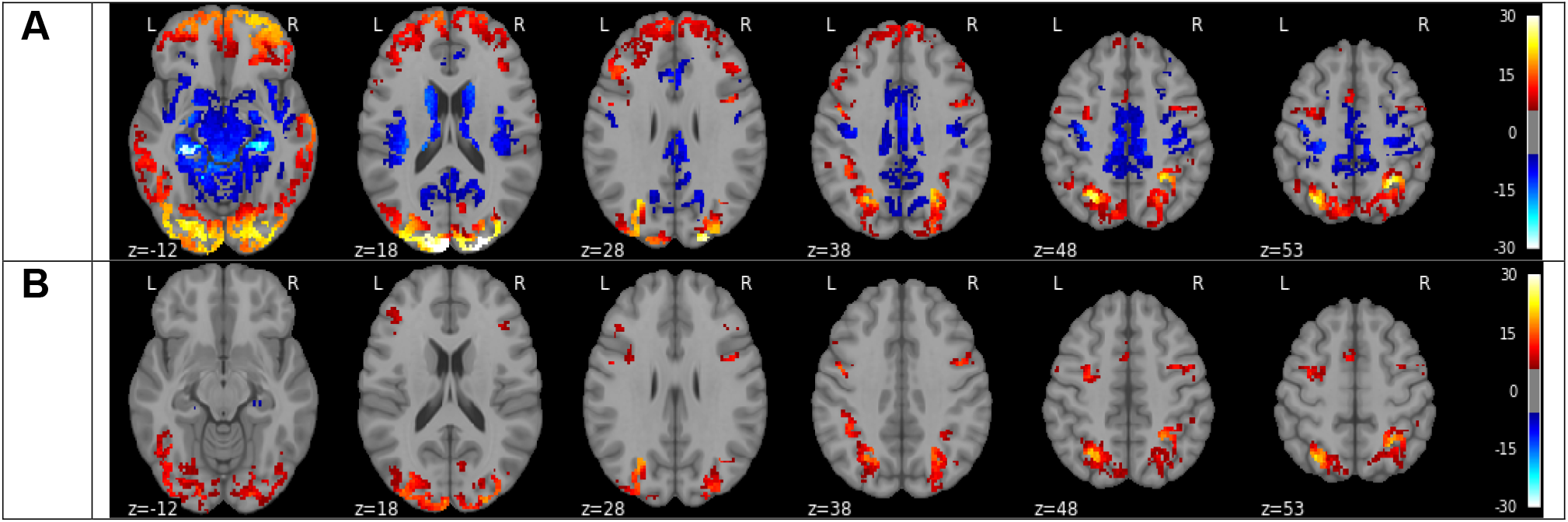
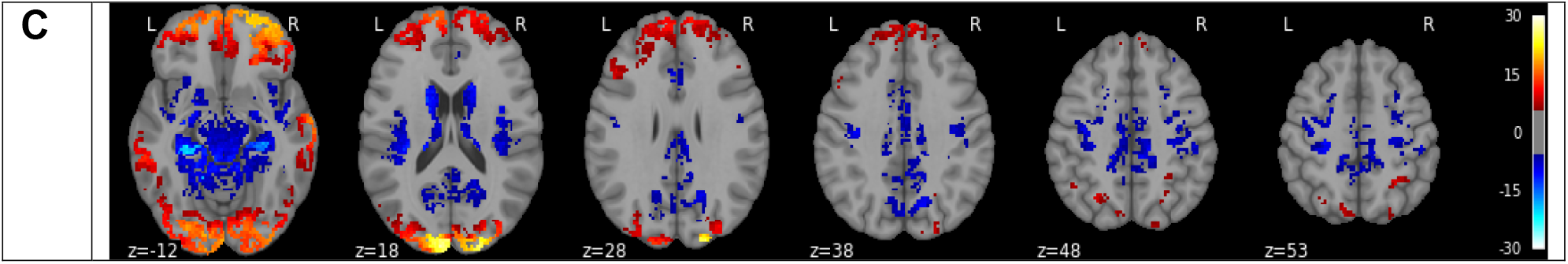
Regional BEN changes induced by the relational task. A) resting BEN vs BEN of the task relational; B) control BEN (BEN of the task control condition) vs BEN of the task relational condition; C) resting BEN minus control BEN. Hot and blue color mean higher and lower BEN at the resting state, respectively. Color bars indicate the display window for the t-value of the paired-t test.

Figure 4 shows the statistically significant BEN difference between the resting state and working memory task performance state. Overall, the three comparison results are highly similar to those found in the relational task vs resting state and control state comparisons (shown in Fig. 3).

**Figure 4:**
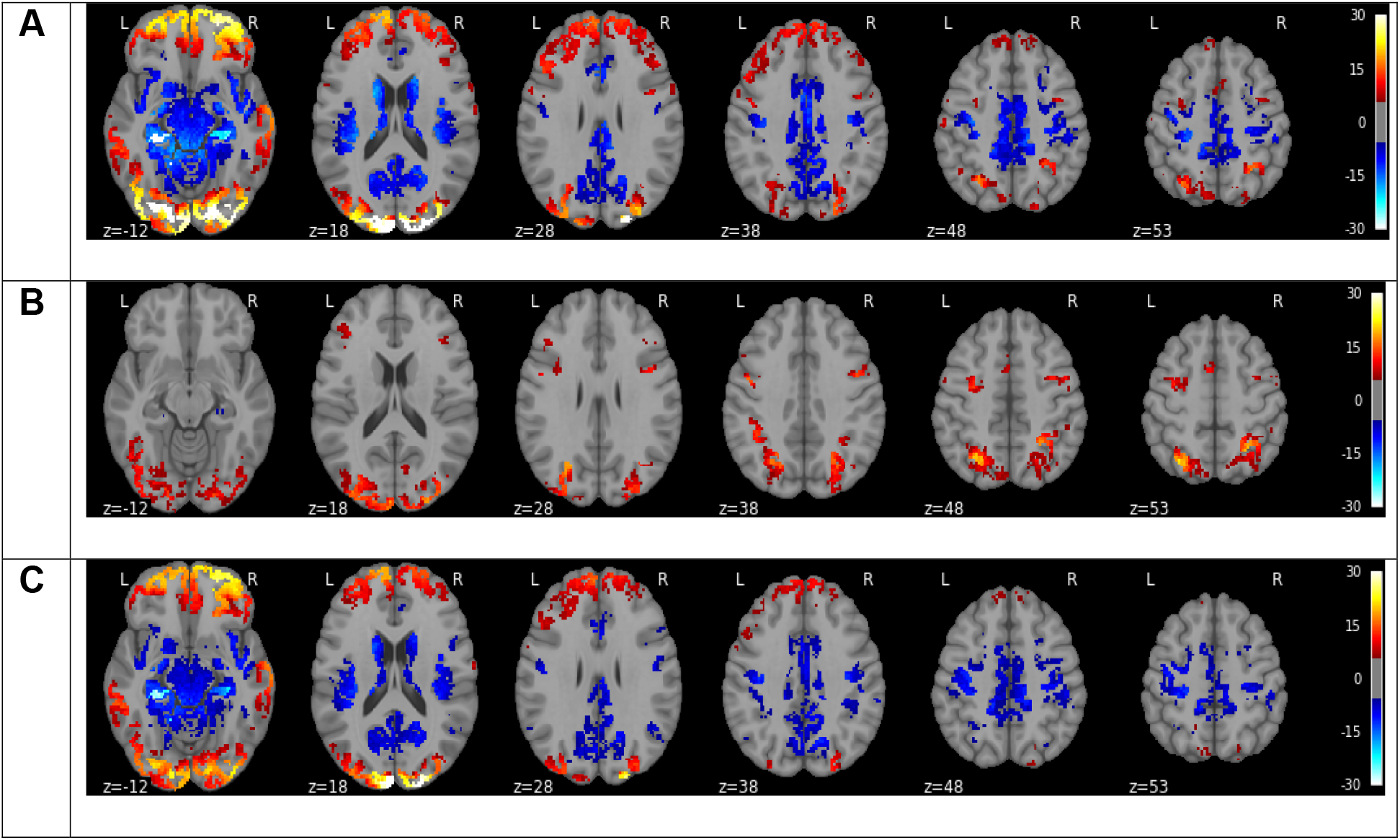
Statistical BEN difference between resting state and working memory task performance. A) resting BEN vs BEN of the task working memory; B) control BEN vs BEN of the task of working memory condition. C) resting BEN minus control BEN (BEN of the non-task condition). Hot and blue color mean higher and lower BEN at the resting state, respectively. Color bars indicate the display window for the t-value of the paired-t test.

Figure 5 shows the statistically significant BEN difference between the resting state and gambling task performance state. These results are very similar to those in Fig. 3-4 though less significant resting BEN vs gambling task BEN was found in OFC and visual cortex (Fig. 5A) relative to those shown in the subfigure A in Fig. 3-4. Compared to the social task (Fig. 2B), relation task (Fig. 3B), and working memory (Fig. 4B), the control BEN vs gambling task BEN difference (Fig. 5B) is less extended in visual cortex, DLPFC, and angular gyrus.

**Figure 5:**
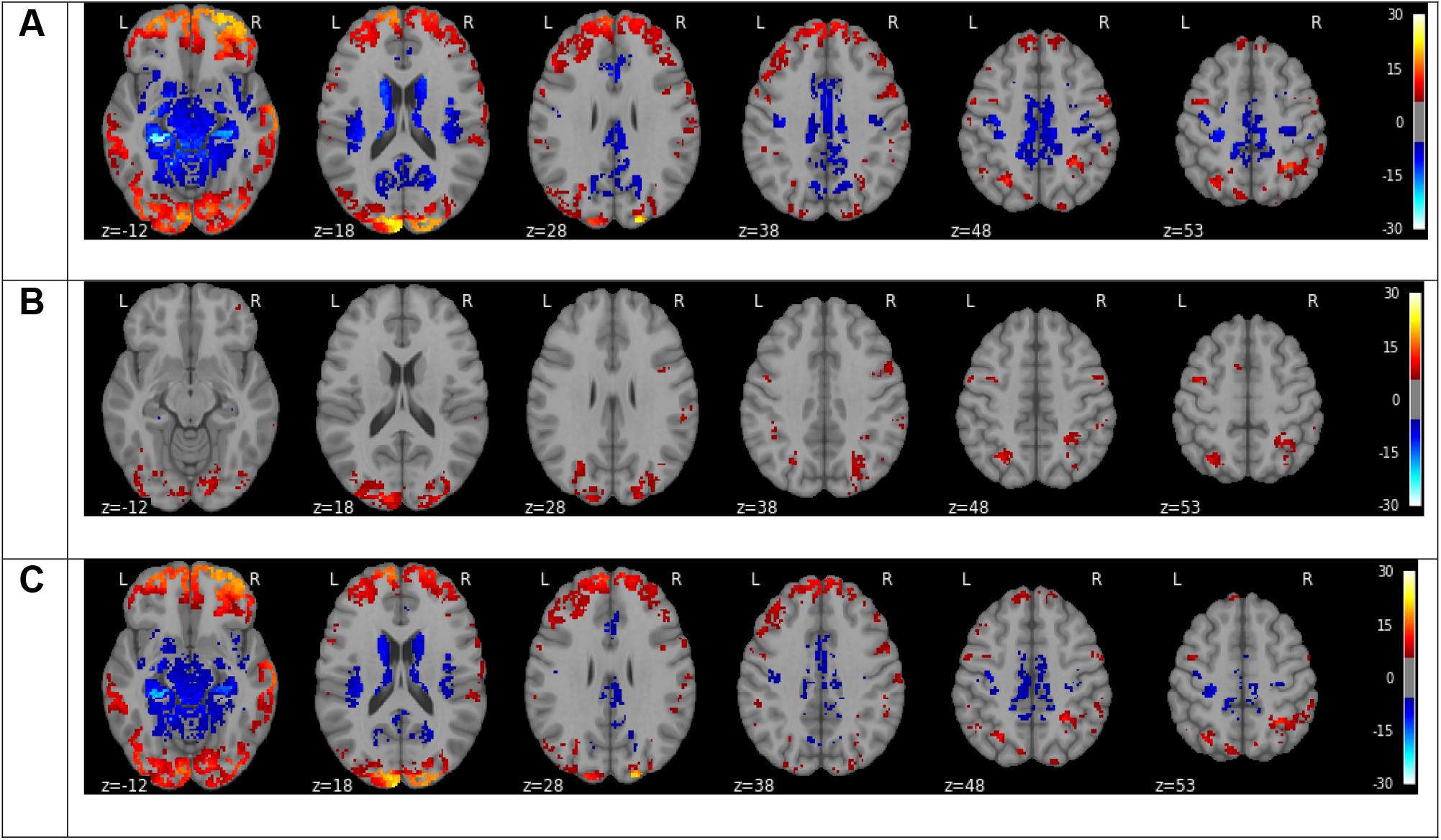
Statistical BEN difference between resting state and gambling task performance. A) resting BEN vs BEN of the task gambling; B) control BEN vs BEN of the task of gambling condition. C) resting BEN minus control BEN (BEN of the non-task condition). Hot and blue color mean higher and lower BEN at the resting state, respectively. Color bars indicate the display window for the t-value of the paired-t test.

Figure 6 shows the statistically significant BEN difference between the resting state and language task performance state. Compared to the resting state (Fig. 6A), language task produced the peripheral BEN reduction and centric BEN increase patterns similar to those reported above in other task fMRI. Compared to the control condition, language task condition showed entropy reduction (hot spots in Fig. 6B) in prefrontal cortex, visual cortex, temporal cortex, and parietal cortex. Compared to the control vs task differences shown in other fMRI as displayed above in Figs. 2-5, the control vs language task difference patterns were very different with bigger cluster in medial and lateral OFC and no difference in DLPFC. The resting vs control comparison (Fig. 6C) showed entropy reduction in regions similar to those found in the control vs task comparison results (Fig. 6B) except for anterior superior prefrontal cortex, DLPFC, hippocamps, and temporal cortex.

**Figure 6:**
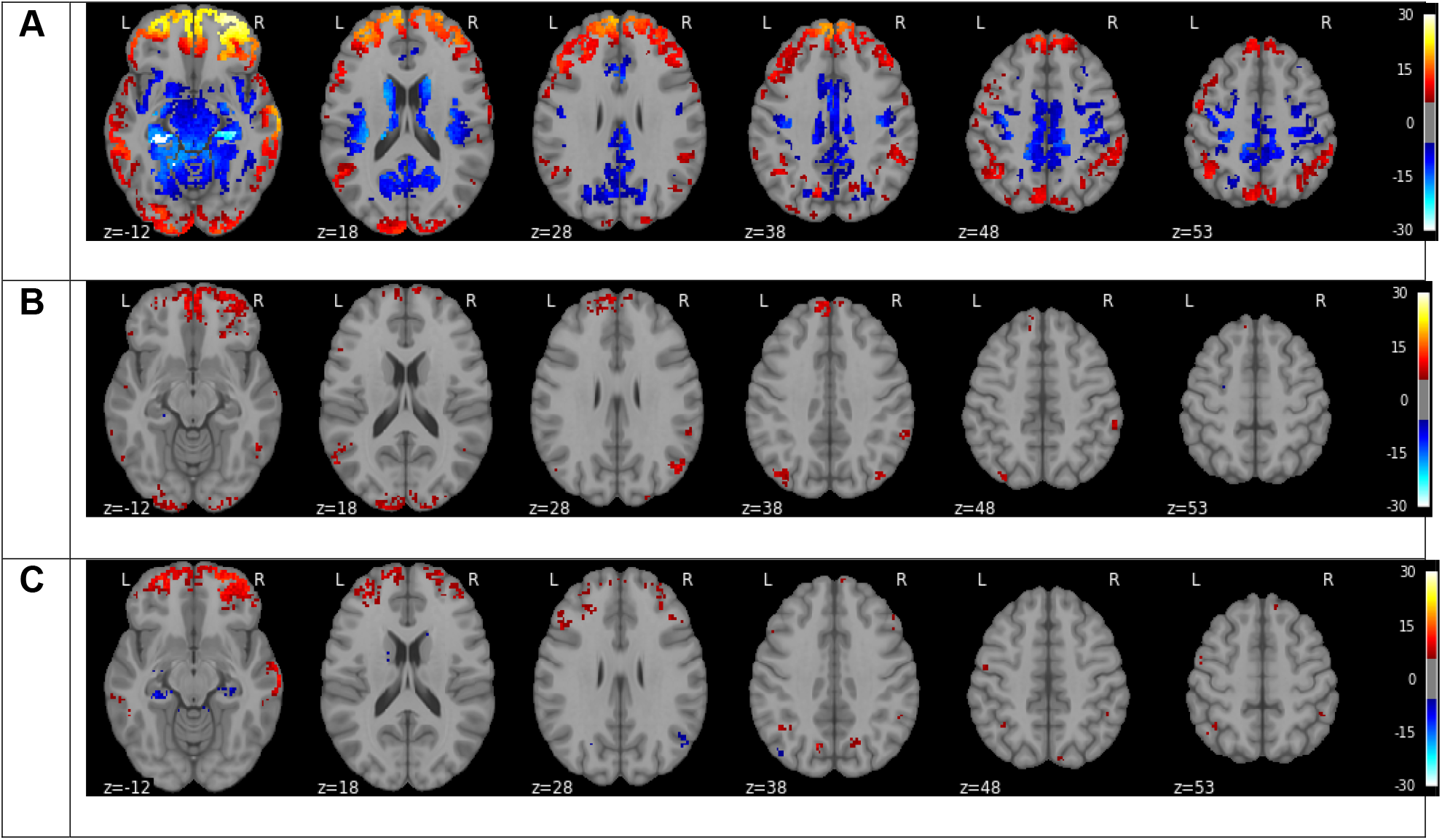
Statistical BEN difference between resting state and language task performance. A) resting BEN vs BEN of the task language; B) control BEN vs BEN of the language task condition; C) resting BEN minus control BEN (BEN of the non-task condition). Hot and blue color mean higher and lower BEN at the resting state or the control state, respectively. Color bars indicate the display window for the t-value of the paired-t test.

Figure 7 shows the statistical analysis results of the emotional task fMRI. The results are similar to those of the language fMRI except for two noticeable spatial pattern difference. In the emotional task fMRI, all three BEN comparison results: resting BEN vs emotion task BEN (Fig. 7A), control BEN vs task BEN (Fig. 7B), and resting BEN vs control BEN (Fig. 7C) showed statistically less significant results in OFC but more significant results in visual cortex compared to those in Fig. 6 (the language task fMRI).

**Figure 7:**
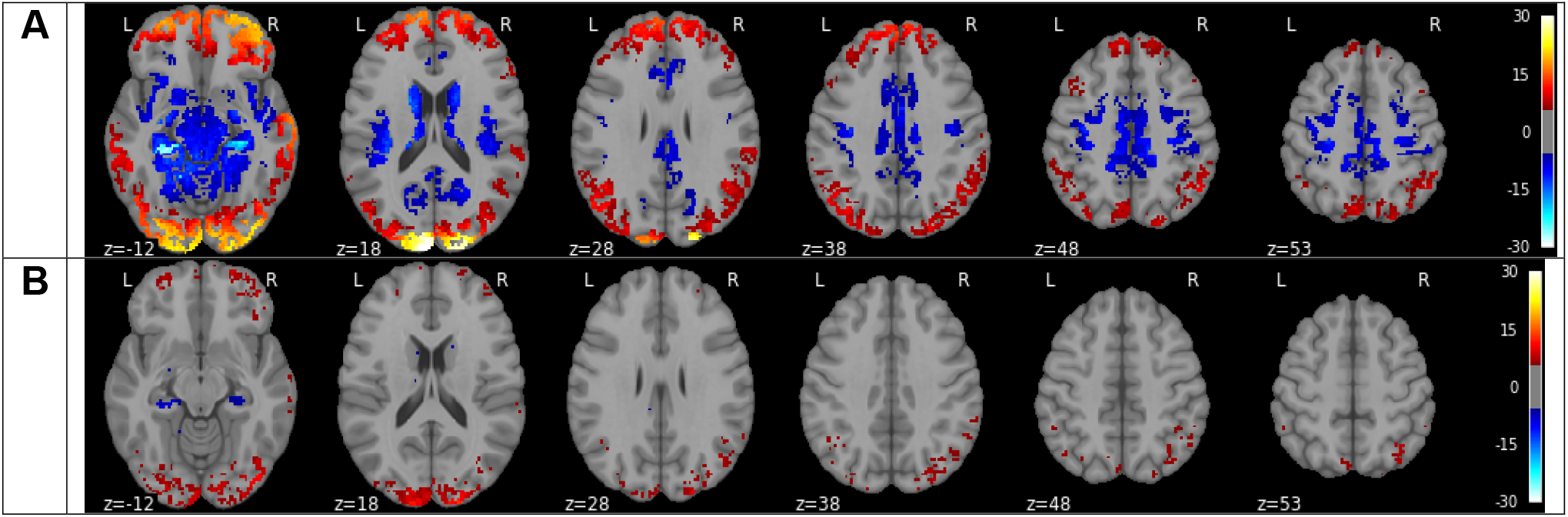
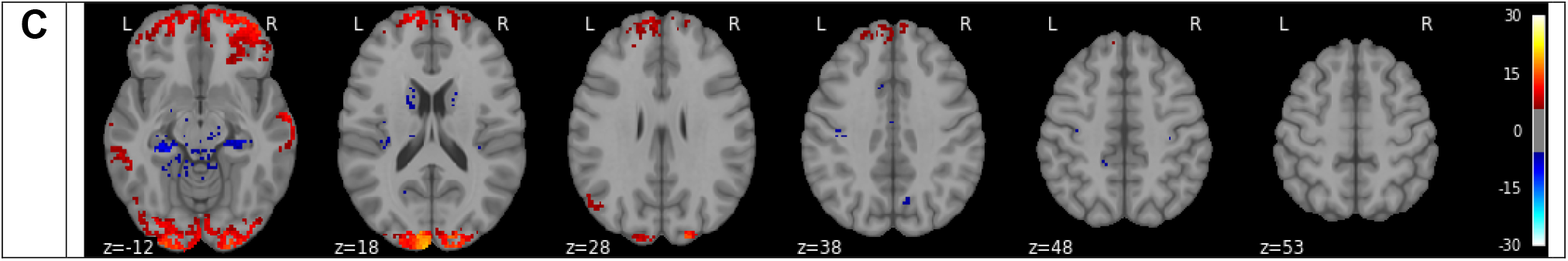
Statistical BEN difference between resting state and emotion task performance. A) resting BEN vs BEN of the task emotion; B) control BEN vs BEN of the task of emotion condition; C) resting BEN minus control BEN (BEN of the non-task condition). Hot and blue color mean higher and lower BEN at the resting state, respectively. Color bars indicate the display window for the t-value of the paired-t test.

## Discussion

We examined task performance induced BEN changes using seven different task Fmri data from HCP. Our major findings are: 1) compared to the resting state, task induced similar BEN change patterns with a clear bifurcation between the peripheral cortical area and the interior cortical and subcortical regions: the peripheral areas showed BEN reduction while the interior brain showed BEN increase; 2) compared to the task control condition, motor task presented similar BEN decrease and increase to the difference between task and resting state. By contrast, the other tasks only showed BEN reduction in task related brain regions; 3) except for the language and emotion task fMRI, the task control condition showed substantial BEN changes compared to the resting state, suggesting a substantial residual task activation in the task control condition though no overt task was performed during that time. Residual task effects were still observed in the control condition in language and emotion task fMRI.

Reduced entropy during task performance in the peripheral cortical areas mainly located in the visual cortex, default mode network(Raichle et al., 2001), and attention network(Damoiseaux et al., 2006), indicating a more regular brain activity in these regions during task performance. This regularity or organization increase is consistent with our previous finding of lower resting entropy in these regions corresponding to better cognitive functions (Wang, 2021) and stronger task activation and deactivations (Liandong Lin et al., 2022). Compared to the resting state, task performance increased BEN mainly in the centric sensorimotor and perception system. High entropy means high irregularity and high information capacity(Shannon, 1948). The increased entropy in the sensorimotor network and perception networks may reflect a need of higher information capacity to allow the brain to have more flexibility to adjust the actions in response to the unknown task (all tasks need button pressing based on the corresponding conditions the subjects saw on the screen).

The control vs task BEN comparison is similar to a standard contrast analysis used in fMRI to detect the task specific brain activations. The difference is that standard contrast analysis is based on mean difference between two contrast conditions while the BEN comparison is to detect the difference of the mean irregularity of two different conditions. Our results showed task specific BEN reductions in all but the motor task. The motor task induced BEN increase in the superior part of the motor network and the limbic sensorimotor system. The other tasks are designed to activate cortical regions involved in high-order functions such as working memory, decision making, emotion etc. BEN reduction in the brain regions underlying these functions suggests that a more organized brain activity is needed in order to execute these high order functions.

The resting state BEN vs task control BEN comparison results showed substantial residual task effects on BEN in the task-free control condition for five of the seven tasks. Emotion and language task fMRI showed the least residual effects in the control condition. This residual effect difference may reflect the different work load of the different fMRI tasks.

A previous study by Nezafati et al. (Nezafati et al., 2020) reported reduced entropy during working memory task in both cortical and subcortical regions, which seems to be contradictory to our findings in the centric brain regions. Their results were based on 100 HCP subjects’ data and they did not report statistical significance level for that reduction. We should note that the HCP rsfMRI data have 1200 data points which is much longer than the task fMRI. In theory, for a data length of around 1200, longer time series tends to produce lower sample entropy value because of the increased number of time segments for the matching process. This has been demonstrated using synthetic data in (Wang et al., 2014). To avoid this confound, we specifically controlled the data length by chopping the longer time series to be the same as the shorter one before calculating BEN. In (Nezafati et al., 2020), task BEN was calculated from the entire task fMRI including the task control condition. Including the control condition may reduce the overall entropy due to the modulation of the systematic on-off task design. To avoid the systematic influence of the low frequency periodic design function, we instead calculated task BEN from the task blocks only after excluding the task control condition from the entire fMRI time series. Nevertheless, we still found similar bidirectional BEN change patterns in the resting BEN vs task BEN comparisons even without excluding the control condition before we calculated the task BEN (data not shown).

Compared to the standard general linear model (GLM) based activation detection results (Barch et al., 2013), our control vs task BEN difference was spatially less extended. This detection sensitivity difference may be attributed to the fundamental difference between magnitude and entropy. GLM is designed to detect the change of the mean signal, while our study was to detect the change of signal irregularity or equivalently the change of the signal change. While this sensitivity difference indicates that mean activity changes induced by task are more sensitive and reliable to detect than the mean change of signal irregularity, our results showed that task did change BEN as we expected. Meanwhile, they suggest that task performance induced brain activity changes are multifaceted.

## Conclusion

Regional BEN is sensitive to task performance in task fMRI.

## Data Availability

Data will be available upon request.

## Acknowledgments

Both imaging and behavior data were provided by the Human Connectome Project, WU-Minn Consortium (Principal Investigators: David Van Essen and Kamil Ugurbil; 1U54MH091657) funded by the 16 NIH Institutes and Centers that support the NIH Blueprint for Neuroscience Research; and by the McDonnell Center for Systems Neuroscience at Washington University in St. Louis. Data and/or research tools used in the preparation of this manuscript were obtained from the National Institute of Mental Health (NIMH) Data Archive (NDA). NDA is a collaborative informatics system created by the National Institutes of Health to provide a national resource to support and accelerate research in mental health.

Dataset identifier(s): [NIMH Data Archive Digital Object Identifier 10.15154/1526336]. This manuscript reflects the views of the authors and may not reflect the opinions or views of the NIH or of the Submitters submitting original data to NDA. The authors thank the Human Connectome Project for open access to its data.

